# Commercial Cannabis Product Testing: Fidelity to Labels and Regulations

**DOI:** 10.1101/2025.03.14.25323943

**Authors:** Sarah Limbacher, Suneeta Godbole, Julia Wrobel, Duncan I. Mackie, Stephen Goldman, Ashley Brooks-Russell

## Abstract

**Background:** In Colorado, regulations for recreational and medical cannabis sales require Tetrahydrocannabinol (THC) concentration is printed on all products. Labeled THC concentrations can vary by +/-15% of what is in the product. Studies show THC concentrations recorded on product labels are not always reflective of the THC concentration in the cannabis product and there is evidence consumers make purchasing decisions based on label claims.

**Aims:** Explore the accuracy of cannabis product labels and differences between THC label accuracy and product type.

**Design:** Data for this analysis come from a larger observational study of cannabis impairment. N=74 flower, concentrate, and edible product samples from licensed Colorado dispensaries were collected and independently tested for THC concentration.

**Setting:** This study was conducted in Colorado, in the Denver Metro Area.

**Participants:** Participants in the study voluntarily enrolled and provided one-gram samples of the cannabis they consumed during the study to be independently tested. The cannabis tested for this analysis was donated on a voluntary basis, not all participants chose to donate.

**Measurement:** The main outcomes of interest for this analysis are accuracy of cannabis product labels compared to observed THC content, accuracy in the context of legally allowable variation, and difference between accuracy by product.

**Findings:** Overall, label values were higher than observed values in flower and edible products (p < 0.001) but was not significant for concentrates (p = 0.85). Flower products were observed to be significantly lower on labels versus the 15% legally allowable range (p = 0.04). Concentrate and edible products were not significantly different (p = 0.9 and p = 0.5, respectively).

**Conclusions:** There is tension between legally allowable THC concentration claims on cannabis product labels and how consumers purchase cannabis. As cannabis policy evolves, standards and regulations that ensure accurate THC concentrations are reported on product labels are urgently needed.

## Introduction

When cannabis was originally scheduled by the U.S. Drug Enforcement Agency (DEA) and the Food and Drug Administration in 1970, its procurement, use, and sale were heavily associated with illicit use (1). Cannabis was first legalized for restricted medical use by the state of Colorado 30 years later, in 2000. Since then, U.S. state-level cannabis legality has boomed, with 38 states legalizing cannabis for medical use, 23 states legalizing recreational use, and 9 states starting ‘low tetrahydrocannabinol (THC)’ programs. As of March 2024, cannabis was recommended to be federally re-scheduled from a Schedule I drug to a Schedule III drug (2).

Despite this widespread legalization in the U.S., regulation of commercially available cannabis products is not universal, with variation between states regarding available products, labeling requirements, and consumer education (3). Because of its historical, federally illegal status, cannabis has not been regulated by the Food and Drug Administration (FDA). Although the FDA has approved one cannabis-derived drug and three cannabis-related drug products, the agency has not approved a “marketing application” for cannabis to treat illness and does not regulate the drug (4). The FDA regulates label requirements of legal drugs, and in lieu of federal regulations, cannabis label policies fall to the discretion of the states (5). The evolution of THC concentration testing and labeling regulations has lagged behind the rapidly growing market and product types, and there are perpetual concerns about product regulation and safety (6). Of concern to state regulators and consumers alike is the accuracy of product labels, including the concentration of THC, the primary psychoactive component.

Label accuracy is an element of product regulation that refers to how well a label represents the content of the product (i.e., accurate representation of THC content). A 2022 study found that there is a lack of uniform labeling practices across states with legal cannabis (5). The same study found that, on average, states have 16.7 different required label attributes and that there is no relationship between years of legal status and labeling requirements. Uniformity in labeling guidelines (as with food labels) promotes awareness of product content and how labels may be interpreted.

One aspect of labeling is THC concentration and the associated permitted variability in concentration. In Colorado, the regulations for label claims for any cannabinoid that has a 2.5 milligram serving or more may be within plus or minus 15% of the concentration actually contained in the product (i.e., cannabis flower that is labeled as 30% THC may actually contain 25.5% to 34.5%). Products with less than 2.5 milligrams per serving may be within +/- 40% of what is reported on the label (7). These regulations were based on plausible variations in concentration, with consideration of other relevant industries and laboratory testing procedures. Other states’ regulations on legally allowable concentration variation are informative. For example, the state of California requires labels to be within +/- 10% of the observed THC concentration (8). Massachusetts’s law states that cannabis labels are required to display “the amount of delta-nine-tetrahydrocannabinol (Δ9-THC) in the package and in each serving” of a marijuana product as expressed in absolute terms and as a percentage of volume (9).

Limited empirical research has reported independent test results of the THC concentration in products available in the commercial marketplace. In a study of the accuracy of cannabis flower labels in Colorado, researchers tested a sample of N=23 different products and found that 78% of the products were over-labeled (i.e., labeled as having an average of 15% higher THC concentration than was found in independent testing) (6). In a study of label accuracy of cannabis edibles from California and Washington, Vandrey et al. also found that 60% of products in their study were labeled as having higher THC content than was observed in the product (10). These studies show that product labels may consistently inflate the actual THC concentration in the product (6,10). This is important because data also show that people rely on reported concentration for purchasing and dosing decisions (11). A major gap in this priori studies is the absence of testing extracted or concentrate products, including edibles, oils, and concentrates (e.g., dabs, shatter, wax, resin), which make up 30% of the legal market (12,13).

The aim of this study was to determine the accuracy of cannabis product labels with respect to THC content among a diverse set of samples of cannabis products (e.g., flower, edibles, and concentrates) purchased in Colorado retail or medical dispensaries as part of a larger study. We explored THC accuracy in the context of product labels and Colorado state regulations. Further, we explore differences between in accuracy by product type.

## Materials and Methods

### Data Collection

From February 20^th^ 2023 to May 6^th^ 2024, cannabis product samples were collected as part of a larger study on the acute effects of cannabis use. Participants in the study provided written consent and during that time were asked if they were willing to bring in the cannabis products that they typically use and were asked if they would provide one gram for independent THC concentration testing. These products were purchased from legal dispensaries in Colorado, and participants were asked to bring the products in their original packaging.

Details of the product label were recorded by research staff, including the following: THC and CBD concentration, batch number, date of sale, dispensary information, and other relevant license numbers. During informed consent, participants were asked if they were willing to donate a sample of approximately one gram of the cannabis they brought in to be independently tested for THC concentration. Cannabis sample donations were weighed, packaged, and placed into a locked safe by participants. For edibles, this typically consisted of 1 gummy, and for participants who vape, this typically consisted of a full cartridge for a vape pen. A third party contracted by our research team retrieved the samples from the research site on the day of donation/data collection. Next, the sample was entered into the METRC system for tracking and then stored at minus 5 degrees Celsius. Periodically, samples were transported to a state-certified laboratory for testing. The Colorado Multiple Institutional Review Board approved study procedures.

### Cannabis Sample Testing

Concentration testing was performed using high-performance liquid chromatography with a diode-array detection (HPLC-DAD) system. All testing and procedures were validated according to ICH Q2 guidelines, and all procedures were audited previously by the Colorado Department of Health and Environment (CDPHE) as well as the Marijuana Enforcement Division (MED). Samples were separated in the Laboratory Information Management System (LIMS) by the license they originated from and the manifest number.

HPLC methodologies were utilized to separate complex mixtures of discrete compounds into their individual populations and quantify the amount of each. Cannabinoids are non-polar molecules and are completely soluble in alcohol. A 30-minute incubation in extraction solution is sufficient to fully extract them. Filtered (0.2 µm) samples (if needed) were processed by HPLC with any validated gradient program. Samples were dissolved in the mobile phase and pumped into any approved reverse-phased C18 column. Sample components separate based on size, polarity, hydrophobicity, and combinations thereof. As compounds elute from the column at specific retention times, they are passed through the diode array detector, which emits a signal proportional to the concentration of the individual components of the sample according to Beer’s law, producing reported observed levels of THC metabolites.

### Statistical Analysis

Statistical analyses were performed using R (version 4.4.1). Given that the data were not normally distributed, the Wilcoxon signed-rank test, a non-parametric alternative to the paired t-test, was employed to assess differences between sample THC values and what was reported on the label. Differences between label and observed values in the context of legally allowable variation were also tested. Results are presented as median values with interquartile ranges (IQRs), and a significance level of α = 0.05 was used to determine statistical significance.

## Results

### Cannabis Characteristics

A total of 74 samples were provided by study participants and independently tested (Figure 1). Products tested include edibles (i.e., gummies, powders; n=29), flower products (i.e., whole flower, joints; n=35), and concentrate products in an oil or solid form (i.e., vaporizer cartridge, ‘shatter’; n=10).

**Figure 1.**
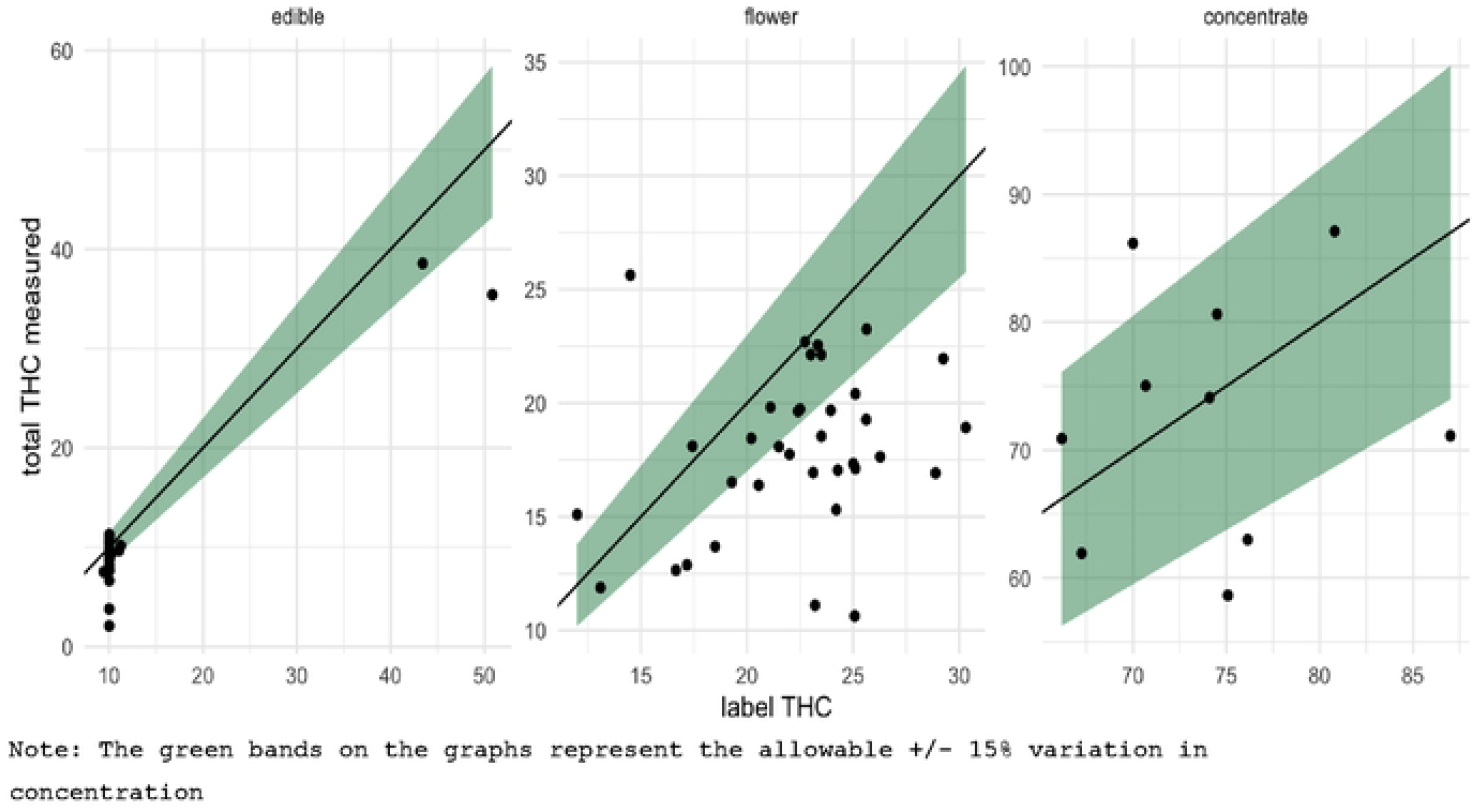
Observed THC product concentration in the context of legally allowable variation.

### Observed THC vs. Labeled THC

We compared THC concentrations reported on labels to observed THC concentrations from laboratory testing by product type. As shown in Table 1, concentrate products had observed THC concentrations that were not significantly different from the label claims (p=.84). Although not statistically significant, the concentrate products had a difference of 6.2% THC between label claims and observed THC concentration, with label claims being greater than observed concentration. The median observed label claim for concentrate products was 74.2% THC concentration as compared to 72.6% for the laboratory-observed THC concentration.

**Table 1:** Difference between label and observed THC concentration by product type.

| Product Type (n= 74) | Labeled THC<br>Median (95% CI) | Observed THC<br>Median (95% CI) | Absolute Difference<br>Median (95% CI) | p | p (+/-15%) |
| --- | --- | --- | --- | --- | --- |
| Edible (n=29) | 10.0 (10.0, 10.0) | 9.13 (7.85, 10.36) | 1.3 (1, 2.3) | <0.001 | 0.54 |
| Flower (n= 35) | 23.12 (20.38, 25.04) | 18.08 (16.46, 19.77) | 4.3 (2.6, 7.5) | <.001 | 0.005 |
| Concentrate (n=10) | 74.3 (70.17, 75.88) | 72.61 (64.96, 79.22) | 6.2 (4.9, 15.2) | 0.85 | 0.9 |

Note. The first test compares the label to the observed THC concentration. The second test compared the observed THC concentration to +/- 15% state, allowed variability in labeled concentration.

Flower and edible products had label claims that were significantly different than the values observed in testing for this study (p<.001 and p<.001, respectively) (Table 1.). In flower products, there was an absolute value difference of 4.3% between label claims and observed THC concentration, with label claims being greater than the observed concentration. Median label claims for flower products were 23.1% THC concentration as compared to the median observed THC concentration of 18.1%.

For edibles, there was a difference of 1.3mg between label claims and observed THC concentration in edible products, with label claims being greater than the observed concentration. The median observed label claim for edible products was 9.3mg THC, as compared to the median label claim of 10mg THC (Table 1).

### Observed THC and State Regulations

We further tested if label claims were significantly different from the +/-15% variation permitted by Colorado law. Overall, concentrate and edible products were not significantly different than allowable variation (p=.9 and p=.5, respectively) (Table 1.). Only flower products had observed THC concentrations that were significantly different (p=.004; Table 1).

These results are further illustrated by Figure 1, which shows THC concentrations by product type and includes shading that represents the permitted +/-15% variation.

## Discussion

This study independently tested a range of cannabis products purchased from cannabis dispensaries in Colorado. This study is consistent with the limited studies that have examined this, which have been conducted in several different states, that find cannabis flower and edible products may not be accurately labeled and do not always reflect observed THC concentrations (6,10). Corroboration of previous findings is significant and suggests that THC concentration in cannabis products is inaccurately marketed across a number of products. Our findings extend prior research by including extracted and concentrate products (e.g., edibles, vape cartridges, shatter) and included edibles that are reflective of the current market, which are widely available, and make up a large component of the market (14).

Although concentrate products are sometimes discussed with public health concerns, such as the potential for greater adverse events associated with the potential for a greater dose, perhaps an unexpected positive side effect of the extraction process is greater accuracy of product labeling.

Label accuracy may be a particularly relevant concern for individuals using cannabis for medical purposes, in which the accuracy of labeling to produce desired and therapeutic effects is a driving motivation for choice of product. There is evidence that people who use cannabis for medical symptom management may purchase products from the recreational or the medical market (15). These individuals may purchase from either retail or medical sources when both options are available in their state, and thus, product label accuracy is a priority regardless of medical or retail status (15). This should be of interest to policymakers because medical cannabis use is prevalent in the United States, an estimated 3.8 million people are registered as medical cannabis patients (16).

There are significant challenges to product testing, particularly with flower products. The inherent variability in a plant means there will be variability within and between flower buds produced by a plant and its clone. In Colorado, state regulations dictate how flower buds are sampled (e.g., 5 samples must be taken totaling 2.5 grams of total mass for harvest batches smaller than one pound) and that these samples are combined and tested as a single test sample representative of the entire pound of material (17).

### Limitations

There are limitations of the study that should be noted. First, there may be some degradation in THC concentration the longer the flower product is stored after harvest (18). Flower products, in particular, may have degraded in concentration related to the length of storage or storage conditions, biasing the results toward larger differences in labeling versus testing. This is not only a limitation of our study but an overall challenge for flower product testing and labeling. Limited information about the duration participants had products before collection is a limitation of the study. Another limitation is that we did not systematically sample dispensaries or products. Instead, we tested products provided by participants for the purposes of a larger research study focused on cannabis impairment. As a result, our findings may not be generalizable to the Colorado dispensary market.

## Conclusion

There are no universal guidelines for cannabis labeling requirements and variability in state testing and label reporting. This is of concern based on consumer and regulator interest in THC concentrations. We found that across product types (flower, edibles, concentrates), there were differences between labeled and lab-tested THC concentrations, with a pattern that packages reported higher THC levels than were found in products. This systematic pattern of over-labeling cannabis products found by our study and others suggest that differences between product labels and observed THC may not just be biological variability. Rather, they reflect systematic forces and incentives in the marketplace, which lead to over-labeling. With rescheduling on the horizon, it is imperative to strategize ways to incentivize federal regulators (like the FDA) to create cannabis label requirements. In lieu of federal regulation, it is necessary to engage state-level regulators to facilitate the evolution of policy that requires cannabis labels to be reliable and accurate.

## Data Availability

All relevant data are within the manuscript and its Supporting Information files.

## Acknowledgements

We would like to acknowledge and thank all staff and participants who made this study possible.

## Primary funding

We would like to acknowledge the funding for this project from the National Institute on Drug Abuse (NIDA R01DA049800).

## References

1. Hudak J. Marijuanas racist history shows the need for comprehensive drug reform. 2020 [cited 2024 Sep 3]; Available from: https://policycommons.net/artifacts/4139172/marijuanas-racist-history-shows-the-need-for-comprehensive-drug-reform/4947315/

2. Federal Register [Internet]. 2024 [cited 2024 Sep 4]. Schedules of Controlled Substances: Rescheduling of Marijuana. Available from: https://www.federalregister.gov/documents/2024/08/29/2024-19370/schedules-of-controlled-substances-rescheduling-of-marijuana

3. Jikomes N, Zoorob M. The Cannabinoid Content of Legal Cannabis in Washington State Varies Systematically Across Testing Facilities and Popular Consumer Products. Sci Rep [Internet]. 2018 Mar 14 [cited 2024 Sep 3];8(1):4519. Available from: https://www.nature.com/articles/s41598-018-22755-2

4. Commissioner O of the. FDA Regulation of Cannabis and Cannabis-Derived Products, Including Cannabidiol (CBD). FDA [Internet]. 2024 Aug 9 [cited 2024 Sep 3]; Available from: https://www.fda.gov/news-events/public-health-focus/fda-regulation-cannabis-and-cannabis-derived-products-including-cannabidiol-cbd

5. Kruger DJ, Korach NJ, Kruger JS. Requirements for Cannabis Product Labeling by U.S. State. Cannabis and Cannabinoid Research [Internet]. 2022 Apr 1 [cited 2024 Sep 3];7(2):156–60. Available from: https://www.liebertpub.com/doi/10.1089/can.2020.0079

6. Schwabe AL, Johnson V, Harrelson J, McGlaughlin ME. Uncomfortably high: Testing reveals inflated THC potency on retail Cannabis labels. PLOS ONE [Internet]. 2023 Apr 12 [cited 2024 Sep 3];18(4):e0282396. Available from: https://journals.plos.org/plosone/article?id=10.1371/journal.pone.0282396

7. Marijuana inspection: sampling procedures | Department of Public Health & Environment [Internet]. [cited 2024 Sep 3]. Available from: https://cdphe.colorado.gov/marijuana-sampling-procedures

8. AMG. California Cannabis Labeling Requirements Explained [Internet]. GAIACA. 2021 [cited 2024 Sep 4]. Available from: https://www.gaiaca.com/california-cannabis-labeling-requirements/

9. THC Label Solutions [Internet]. [cited 2024 Sep 4]. Massachusetts Cannabis Package and Label Laws and Requirements. Available from: https://thclabelsolutions.com/blog/massachusetts-cannabis-package-and-label-laws-and-requirements/

10. Vandrey R, Raber JC, Raber ME, Douglass B, Miller C, Bonn-Miller MO. Cannabinoid Dose and Label Accuracy in Edible Medical Cannabis Products. JAMA [Internet]. 2015 Jun 23 [cited 2024 Sep 3];313(24):2491–3. Available from: 10.1001/jama.2015.6613

11. Cash MC, Cunnane K, Fan C, Romero-Sandoval EA. Mapping cannabis potency in medical and recreational programs in the United States. PLoS One. 2020;15(3):e0230167.

12. Colorado Cannabis Prices and Trends | Headset [Internet]. [cited 2024 Sep 3]. Available from: https://www.headset.io/markets/colorado

13. Tableau Public [Internet]. [cited 2024 Oct 23]. Colorado MED Dashboard. Available from: https://public.tableau.com/app/profile/cu.business.research.division/viz/ColoradoMEDDashboard/Overview

14. Google Docs [Internet]. [cited 2024 Oct 23]. MED Quarterly Update Report Q4 Final.pdf. Available from: https://drive.google.com/file/d/1LQ6BUCZYXqZVvEZZWkW9oEEezzSdHZlA/view?usp=sharing&usp=embed_facebook

15. Jameson LE, Conrow KD, Pinkhasova DV, Boulanger HL, Ha H, Jourabchian N, et al. Comparison of State-Level Regulations for Cannabis Contaminants and Implications for Public Health. Environmental Health Perspectives [Internet]. 2022 Sep [cited 2024 Sep 3];130(9):097001. Available from: https://ehp.niehs.nih.gov/doi/full/10.1289/EHP11206

16. Project MP. MPP. [cited 2024 Sep 3]. Map of State Marijuana Laws. Available from: https://www.mpp.org/issues/legalization/map-of-state-marijuana-laws/

17. CDPHE Marijuana Flower Sampling Protocol_Dec 2020 - Google Docs [Internet]. [cited 2024 Oct 23]. Available from: https://docs.google.com/document/d/1eQ3El_ZH65HTru9Czl6-z2SLzTPgGeDyaB6QIgwhweg/edit?tab=t.0

18. Meija J, McRae G, Miles CO, Melanson JE. Thermal stability of cannabinoids in dried cannabis: a kinetic study. Anal Bioanal Chem. 2022 Jan;414(1):377–84.

19. dcc_commercial_cannabis_regulations-1.pdf [Internet]. [cited 2024 Sep 3]. Available from: https://cannabis.ca.gov/wp-content/uploads/sites/2/2024/08/dcc_commercial_cannabis_regulations-1.pdf

